# A comparison between early presentation of dementia with Lewy Bodies, Alzheimer’s disease and Parkinson’s disease: evidence from routine primary care and UK Biobank data

**DOI:** 10.1101/2023.01.04.23284188

**Authors:** Thomas Nedelec, Baptiste Couvy-Duchesne, Aube Darves-Bornoz, Raphaël Couronne, Fleur Monnet, Laurène Gantzer, Béranger Lekens, Yeda Wu, Nicolas Villain, Anette Schrag, Stanley Durrleman, Jean-Christophe Corvol

## Abstract

**Objective:** To simultaneously contrast prediagnostic clinical characteristics of individuals with a final diagnosis of dementia with Lewy Bodies, Parkinson’s disease, Alzheimer’s disease compared to controls without neurodegenerative disorders.

**Methods:** Using the longitudinal THIN database in the UK, we tested the association of each neurodegenerative disorder with a selected list of symptoms and broad families of treatments, and compared the associations between disorders to detect disease-specific effects. We replicated the main findings in the UK Biobank.

**Results:** We used data of 28,222 patients with PD, 20,214 with AD, 4,682 with DLB and 20,214 controls. All neurodegenerative disorders were significantly associated with the presence of multiple clinical characteristics before their diagnosis including sleep disorders, falls, psychiatric symptoms and autonomic dysfunctions. When comparing DLB patients with patients with PD and AD patients, falls, psychiatric symptoms and autonomic dysfunction were all more strongly associated with DLB in the five years preceding the first neurodegenerative diagnosis. The use of statins was lower in patients who developed PD and higher in patients who developed DLB compared to AD. In PD patients, the use of statins was associated with the development of dementia in the five years following PD diagnosis.

**Interpretation:** Prediagnostic presentations of falls, psychiatric symptoms and autonomic dysfunctions were more strongly associated with DLB than PD and AD. This study also suggests that whilst several associations with medications are similar in neurodegenerative disorders, statin usage is negatively associated with Parkinson’s Disease but positively with DLB and AD as well as development of dementia in PD.

## Introduction

Neurodegenerative diseases represent one of the main public health issues in western societies. In 2019, about 57 million individuals were living with dementia worldwide.^1^ Alzheimer’s disease (AD) accounts for 60-70% of dementia cases (29 million) and represents the first cause of dementia while dementia related to synucleinopathies (Parkinson’s disease dementia and dementia with Lewy bodies) are the second most common neurodegenerative cause of dementia.^2^ Parkinson’s disease (PD) is the second cause of neurodegenerative disease, estimated worldwide to affect 6.1 million individuals in 2016.^3^ All these neurodegenerative diseases are known to have a very long preclinical stage. For AD, accumulation of amyloid can precede a clinical diagnosis of dementia by up to 20 years.^4^ Synucleinopathies, including PD and DLB, also have a long prodromal stage as demonstrated by isolated REM sleep behavioural disorders which can precede symptomatic PD or DLB by 10 years or more ^5^. The first dysautonomic signs have been estimated to start as early as 15 years before motor symptoms in PD.^6,7^

Analyses on general practitioners (GP) databases have already led to the identification of disease-specific prodromal symptoms of PD and AD as well as risk factors in the primary care data^8,9^ However, it is unknown how the prodromal features and risk factors of these disorders differ. Such analyses on dementia with lewy bodies are also missing. Overall, cross-disorder analyses focusing on shared and specific associations with neurodegenerative diseases are rare.^10^ We undertook cross-disorder analyses in order to identify prediagnostic associations and risk factors of the most common neurodegenerative conditions (AD, PD and DLB), using data collected by general practitioners over a long time window. We sought to identify disorders and drug prescriptions associated with AD, PD and DLB, as well as those that are specific to each condition. Predictors of dementia in PD observable in primary care are also missing.^11^ In a secondary analysis, we aimed at understanding which drug prescriptions are associated with the presence of dementia in the five years following a first PD diagnosis. We relied on the Health Improvement Network UK primary care for discovery and the UK Biobank (UKB) for selected replication.

## Methods

### Study design and participants

#### THIN database

We used The Health Improvement Network (THIN®) database,12 a large European standardized database of anonymized Electronic Medical Records collected at the physicians’ level by the company Cegedim and coded using the International Classification of Diseases, 10th Revision (ICD-10) codes. Data were collected in 400 general practices, representing around 6% of the UK population.9 Several reports have shown that electronically coded diagnoses in this database are representative of the UK general practice population in terms of demographics and type of consultation.12,13 For each patient and visit, we obtained, the diagnosis and prescription established during the visit, as well as all ongoing prescriptions and associated diagnoses.

We defined our neurological cases based on ICD10 codes given by the general practitionners of the THIN database recorded between January 1996 and April 2020. For the neurodegenerative disorders of interest, AD cases corresponded to codes F00 and G30, PD to code G20 and dementia with Lewy Bodies to code G31.8 (and READ code Eu025, to exclude frontotemporal dementia). We extracted all DLB patients in the database, and a random subsample of patients with AD or with PD, with at least 2 years of follow-up. We defined age at onset as the age at the first record coded with AD, PD or DLB diagnosis. Controls were screened for any history of Alzheimer’s disease (F00 et G30), Parkinson’s disease (G20), frontotemporal dementia (G31.0 and G31.8), dementia with Lewy Bodies (G31.8), Huntington’s disease (G10), or multiple sclerosis (G35). We had access to 20,214 controls matched with AD cases on sex and age +/- 1 year at last record in the database from a precedent study which was specific to AD.^14^

#### UK Biobank replication sample

For replication, we used the UKB, a large population-based cohort which consists in 502,492 unselected volunteers from the UK, recruited between 2006 to 2010, of which 487,409 underwent genotyping and extensive phenotyping.^15^ Phenotypic data of interest include age, sex, Body Mass Index, townsend deprivation index and the age they completed full time education. Cholesterol data was measured from blood collected in the baseline visit. Informed consent was obtained from all participants registered in the UK Biobank.

In the UKB, we included the 1,005 patients with a reported diagnosis of AD, the 2,268 patients with PD, and the 2,732 patients with a report of dementia (Table 3). The clinical outcomes (Parkinson’s field ID 42030, Alzheimer’s field ID 42020, all dementia field ID 42018) were generated by the UKB (category 42), by crossing UKB baseline self-reports with linked hospital admissions and death register data. We excluded from further analysis the 69 patients who had records of both AD and PD diagnoses. (see eTable 7 and eTable 8 for statistics on this population). Finally, we only considered patients with at at least one UKB visit before a diagnosis of neurodegenerative disease.

#### Health conditions and prescriptions of drugs of interest

We considered symptoms previously reported to be associated with one of these neurodegenerative diseases in the literature and likely to be coded by a GP. We included in the analysis memory problems, tremor, confusion, hallucinations, sleep disorders, constipation, anxiety, depression, falls, hypotension, urinary tract disorders and abnormal weight loss. We used the READ codes and the International Classification of Diseases, 10th Revision (ICD10) to extract health conditions (the mapping is provided in eTable 16). We also conducted analysis on a preselected list of classes of medications previously reported to be associated with one of the neurodegenerative diseases, including all classes of anti-hypertensive medications, laxatives, statins, and medications used for neurological disorders, including treatments against depression, anxiety, and psychosis (eTable 17). We relied on the Anatomical Therapeutic Chemical (ATC) Classification System^16,17^ available directly in the THIN database. In the UKB, we used the medication-use data collected by trained nurses during interviews. The UKB classifies medications into 6,745 categories of which 1,809 were reported by 10 or more people.^16^ We mapped the medication categories into ATC codes (1,752 over 1,809 were mapped) as previously done in a genetic analysis of the UKB.^16^

### Statistical analysis

We first used the THIN database to estimate the association between the preselected list of health conditions or medication classes and each specific neurodegenerative disorder. We estimated the odds ratios using a logistic regression corrected for sex and age at index date. We reported the OR estimates for different time windows prior to the neurodegenerative onset: (0 - 5] years, (5 – 10] years, and (10-15] years, in order to show their progression over time. The index date for each AD-control pair was defined as the first date of AD diagnosis of the corresponding patient with AD. The index date for PD and DLB was defined as age of onset. We corrected for multiple comparison using Bonferroni corrections. This led to using a significance threshold of p < 0.004 (p=0.05 with Bonferroni correction for 12 potential exposures, 2-tailed test). All analyses were conducted using the StatsModel library in Python. Next, we compared the OR between the three neurodegenerative disorders (PD vs. AD, DLB vs. AD and DLB vs. PD). We also compared the association of prediagnostic medication use between PD patients who developed dementia in the five years after a first PD diagnosis and those who did not. For this analysis, we excluded patients with a diagnosis of dementia before a first PD diagnosis. We compared these two groups in terms of medication usage in the [0-5] years before PD diagnosis.

In the UKB, we compared the AD and PD groups to replicate the significant results for all classes of medications considered in the THIN study. We used logistic models and controlled for sex, assessment center, age at diagnosis, age at entry in the cohort, and also for the Townsend deprivation index, body mass index and education level. In a sensitivity analysis, we constrasted PD patients with the group of patients with a report of AD or all field dementia. When testing associations of PD, AD or all field dementia with the class of lipid-modifying agents, we further corrected for Apolipoprotein E (APOE) status (carriers of at least one epsilon 4 variant). We finally compared the associations with cholesterol, the LDL and HDL levels between PD and AD patients.

## Results

In the THIN database, our full initial analytical cohort included 20,214 patients with a final diagnosis of AD, 28,222 patients with a diagnosis of PD, 4,682 patients with a diagnosis of DLB and 20,214 control patients. Table 1 describes the characteristics of the different populations. The median age at diagnosis was younger for PD (77 (70-83) years) than for DLB (80 (74-85) years) and AD patients (82 (76-87) years) and for controls (82 (76-87) years). There were more men than women in the PD and DLB cohorts (sex ratio of 57%) and the opposite in the AD and controls cohorts (sex ratio of 31%). 13,625 AD patients, 14,539 PD patients, 2,279 DLB patients and 10,205 controls had data available for more than 10 years before diagnosis. Demographic characteristics were similar between the different retrospective follow-up intervals (0 to 5, 10 or 15 years before diagnosis, eTable1). Only 5% of AD patients had one prescription of antiparkinsonian drugs while 76% of PD patients had at least two prescriptions of antiparkinsonian drugs (eTable 2).

**Table 1:** Characteristics of patients with Alzheimer’s disease, with Parkinson’s disease, with Dementia with Lewy Bodies and controls. The data shown are numbers (%) and medians (IQR).

|  | Total |  |  |  | With >=5 years of retrospective data |  |  |  |
| --- | --- | --- | --- | --- | --- | --- | --- | --- |
|  | Alzheimer's<br>disease (n=20214) | Parkinson's<br>disease (n=28222) | Dementia with<br>Lewy Bodies<br>(n=4682) | Controls<br>(n=20214) | Alzheimer's<br>disease<br>(n=17871) | Parkinson's<br>disease<br>(n=22281) | Dementia with<br>Lewy Bodies<br>(n=2712) | Controls<br>(n=15361) |
| Sex: Women | 14106 (69%) | 12332 (43%) | 2050 (43%) | 14106 (69%) | 12531 (70%) | 9532 (42%) | 1156 (42%) | 10624 (69%) |
| Men | 6108 (31%) | 15890 (57%) | 2632 (57%) | 6108 (31%) | 5340 (30%) | 12749 (58%) | 1556 (58%) | 4737 (31%) |
| Birth year | 1926 (1921-1933) | 1929 (1921-1938) | 1932 (1926-1938) | 1925 (1918-1933) | 1927 (1921-1933) | 1931 (1923-1940) | 1932 (1927-1939) | 1927 (1920-1934) |
| Age at index date | 82 (76-87) | 77 (70-83) | 80 (74-85) | 82 (76-87) | 82 (76-87) | 77 (70-83) | 79 (74-84) | 82 (76-87) |
| Person-years of data available before index date | 13 (8-18) | 10 (6-16) | 10 (1-18) | 10 (5-17) | 14 (10-18) | 12 (9-18) | 17 (12-21) | 13 (9-18) |
| Number of visits per year | 7 (4-11) | 7 (4-12) | 10 (6-17) | 7 (3-11) | 7 (4-10) | 7 (4-11) | 7 (4-11) | 7 (4-11) |

**Table 2:** OR and 95% CIs for a logistic model assessing the association of at least one prescription in the considered class of treatment in the five years preceding a first Parkinson’s disease diagnosis with a diagnosis of dementia in the 5 years following a first Parkinson’s disease diagnosis. In the multivariable model, we included all seven health conditions. The treatment has to be prescribed in 0–5 years before Parkinson’s disease diagnosis. OR=odds ratio. Analyses are corrected for age at PD diagnosis and sex. P values are corrected for multiple comparisons in the univariable analysis.

|  | Prevalence of at least one prescription in the 5 years before PD diagnosis |  | Univariable analysis |  | Multivariable analysis |  |
| --- | --- | --- | --- | --- | --- | --- |
|  | PD patients with dementia in the five years after PD diagnosis (n= 1682) | PD patients without dementia in the five years after PD diagnosis (n = 6924) | OR (95% CI) | p value | OR (95% CI) | p value |
| Drugs used against hypertension | 1074 (63.85%) | 4126 (59.59%) | 1.01 (0.9,1.14) | 0.805 (5.633) | 0.85 (0.75,0.96) | 0.01 |
| Drugs used against hypercholesterolemia | 579 (34.42%) | 1814 (26.2%) | 1.5 (1.33,1.69) | <0.001 (<0.001) | 1.49 (1.31,1.69) | <0.001 |
| Drugs used against diabetes | 140 (8.32%) | 535 (7.73%) | 1.07 (0.87,1.31) | 0.517 (3.62) | 0.9 (0.73,1.11) | 0.334 |
| Drugs used against constipation | 671 (39.89%) | 1685 (24.34%) | 1.56 (1.39,1.76) | <0.001 (<0.001) | 1.42 (1.25,1.6) | <0.001 |
| Drugs used against depression | 653 (38.82%) | 2212 (31.95%) | 1.56 (1.38,1.75) | <0.001 (<0.001) | 1.44 (1.27,1.63) | <0.001 |
| Drugs used against anxiety | 255 (15.16%) | 1112 (16.06%) | 1.05 (0.9,1.23) | 0.504 (3.53) | 0.85 (0.72,1.0) | 0.044 |
| Drugs used against psychosis | 430 (25.56%) | 1262 (18.23%) | 1.37 (1.2,1.57) | <0.001 (<0.001) | 1.23 (1.08,1.42) | 0.003 |

All conditions were more frequent than in controls in all three neurodegenerative diseases during the period 0-5 years before diagnosis (Figure 1). The prevalence of tremor, sleep disorders, constipation, anxiety and urinary tract disorders was higher in PD and DLB than in controls up to 15 years prior diagnosis, and anxiety and depression only for DLB (eTable3). The pre-diagnosis profile of the 12 health conditions investigated in the THIN study in AD, PD, DLB and controls over time are shown in Figure 2 (eTable4). Treatments against depression, anxiety and constipation were all more frequently prescribed in individuals with a later diagnosis of AD, PD and DLB (Figure 1). A higher frequency of benzodiazepines and serotonin reuptake inhibitors prescription was observed in the years preceding the diagnosis of all three diseases (Table 4). Lower antidiabetic medications use, including insulins, biguanids, sulfonylureas and DDP4 inhibitors, was observed before the diagnosis of AD and PD (Table 4). A lower prescription of agents acting on the renin–angiotensin–aldosterone system and selective beta-2-adrenoreceptor agonists was significantly observed before the diagnosis of the three diseases when compared to controls (Table 4).

**Table 3:** Characteristics of patients with any types of dementia, Alzheimer’s disease and Parkinson’s disease in the UK Biobank. The data shown are numbers (%) or medians (IQR).

|  | All |  |  |  | With at least one visit before diagnosis |  |
| --- | --- | --- | --- | --- | --- | --- |
|  | Total UK Biobank | All dementias | Alzheimer | Parkinson | Alzheimer | Parkinson |
| Total | 502492 | 2732 | 1005 | 2268 | 920 | 1272 |
| Women (n, %) | 273377 (54%) | 1216 (44%) | 491 (48%) | 879 (38%) | 465 (50%) | 505 (39%) |
| Men (n, %) | 229115 (46%) | 1516 (56%) | 514 (52%) | 1389 (62%) | 455 (50%) | 767 (61%) |
| BMI | 26.86 (24.23-29.98) | 27.24 (24.41-30.3) | 26.75 (23.96-29.54) | 27.25 (24.7-30.16) | 26.66 (23.79-29.37) | 27.38 (24.75-30.26) |
| Education level | 16 (15-17) | 15 (15-16) | 15 (15-16) | 16 (15-17) | 15 (15-16) | 16 (15-17) |
| Townsend deprivation | -2.14 (-3.64-0.55) | -1.67 (-3.46-1.71) | -1.99 (-3.62-1.31) | -2.29 (-3.71-0.7) | -2.02 (-3.64-1.24) | -2.17 (-3.67-0.88) |
| Year of birth | 1950 (1945-1958) | 1943 (1940-1946) | 1942 (1940-1946) | 1944 (1941-1948) | 1942 (1940-1946) | 1944 (1941-1948) |
| Age at first visit | 58 (51-64) | 66 (62-68) | 66 (63-68) | 64 (60-67) | 66 (63-68) | 65 (61-68) |
| Age at diagnosis |  | 71 (67-74) | 72 (68-74) | 65 (58-71) | 72 (69-74) | 70 (66-73) |

**Table 4:** Odds ratios corresponding to Alzheimer’s and Parkinson’s disease and Dementia with Lewy Bodies and to the comparison between Parkinson’s and Alzheimer’s disease (0-5] years before a first diagnosis on the THIN database and odds ratios corresponding to the comparison between Parkinson’s and Alzheimer’s disease and between Parkinson’s disease and all dementias with presentations before diagnosis of the corresponding neurodegenerative disorders in the UK Biobank replication sample. P values are corrected for multiple comparisons in the THIN analysis. In the THIN analysis, odds ratios are corrected for age at diagnosis and sex and in the UK Biobank replication sample, for age at diagnosis, age at first visit, sex, center, BMI, education level and townsend deprivation index.** cannot be calculated because an insufficient number of presentations was recorded.

|  | THIN analysis |  |  |  |  |  |  |  | UK Biobank replication sample |  |  |  |
| --- | --- | --- | --- | --- | --- | --- | --- | --- | --- | --- | --- | --- |
|  | Alzheimer's disease |  | Parkinson's disease |  | Lewy Body disease |  | Parkinson's disease vs Alzheimer's disease |  | Parkinson's disease vs Alzheimer's disease |  | Parkinson vs all dementia |  |
|  | OR (95%CI) | p value | OR (95%CI) | p value | OR (95%CI) | p value | OR (95%CI) | p value | OR (95% CI) | P value | OR (95% CI) | P value |
| Diuretics | 0.74 (0.71,0.77) | <0.001 (<0.001) | 0.93 (0.89,0.97) | 0.002 (0.035) | 0.85 (0.78,0.93) | <0.001 (0.006) | 1.25 (1.2,1.31) | <0.001 (<0.001) | 0.88 (0.67, 1.14) | 0.324 | 0.85 (0.68, 1.07) | 0.169 |
| Beta-blocking agents | 0.86 (0.82,0.9) | <0.001 (<0.001) | 1.14 (1.08,1.19) | <0.001 (<0.001) | 1.21 (1.11,1.33) | <0.001 (0.001) | 1.32 (1.26,1.38) | <0.001 (<0.001) | 0.88 (0.68, 1.13) | 0.309 | 0.78 (0.62, 0.97) | 0.028 |
| Calcium-channel blockers | 0.77 (0.73,0.81) | <0.001 (<0.001) | 0.77 (0.73,0.81) | <0.001 (<0.001) | 0.93 (0.85,1.02) | 0.144 (2.441) | 1.01 (0.96,1.07) | 0.566 (9.624) | 0.84 (0.64, 1.09) | 0.18 | 0.69 (0.55, 0.87) | 0.001 |
| Agent acting on angiotensin system | 0.71 (0.68,0.74) | <0.001 (<0.001) | 0.73 (0.7,0.77) | <0.001 (<0.001) | 0.8 (0.73,0.87) | <0.001 (<0.001) | 1.05 (1.0,1.1) | 0.049 (0.826) | 0.93 (0.76, 1.16) | 0.534 | 0.74 (0.62,0.89) | 0.001 |
| Statins | 0.98 (0.93,1.02) | 0.304 (5.175) | 0.78 (0.75,0.82) | <0.001 (<0.001) | 1.38 (1.27,1.51) | <0.001 (<0.001) | 0.82 (0.78,0.86) | <0.001 (<0.001) | 0.77 (0.63, 0.94) | 0.01 | 0.73 (0.62, 0.86) | <0.001 |
| Fibrates | 0.83 (0.65,1.07) | 0.151 (2.565) | 0.81 (0.64,1.04) | 0.094 (1.591) | 0.64 (0.37,1.1) | 0.104 (1.776) | 0.97 (0.76,1.23) | 0.789 (13.421) | 0.47 (0.16, 1.33) | 0.154 | 0.54 (0.21, 1.36) | 0.19 |
| Other lipid modifying agents | 0.83 (0.7,0.99) | 0.041 (0.699) | 0.82 (0.69,0.98) | 0.026 (0.449) | 1.05 (0.75,1.45) | 0.792 (13.457) | 0.95 (0.8,1.14) | 0.579 (9.842) | 1.08 (0.75, 1.56) | 0.679 | 1.02 (0.75, 1.38) | 0.921 |
| Insulins and analogues | 0.72 (0.61,0.85) | <0.001 (0.002) | 0.74 (0.63,0.87) | <0.001 (0.004) | 1.04 (0.78,1.37) | 0.807 (13.721) | 1.02 (0.86,1.21) | 0.828 (14.079) | 0.59 (0.35, 1.0) | 0.051 | 0.49 (0.31, 0.77) | 0.002 |
| Biguanides | 0.77 (0.71,0.84) | <0.001 (<0.001) | 0.77 (0.7,0.83) | <0.001 (<0.001) | 0.89 (0.76,1.05) | 0.161 (2.736) | 0.99 (0.91,1.08) | 0.848 (14.417) | 0.78 (0.53, 1.15) | 0.213 | 0.61 (0.44, 0.85) | 0.003 |
| Sulfonylureas | 0.76 (0.69,0.84) | <0.001 (<0.001) | 0.83 (0.75,0.91) | <0.001 (0.001) | 0.64 (0.52,0.79) | <0.001 (<0.001) | 1.06 (0.96,1.17) | 0.234 (3.985) | 1.21 (0.67, 2.17) | 0.525 | 0.75 (0.47, 1.2) | 0.232 |
| DDP4 inhibitors | 0.62 (0.48,0.79) | <0.001 (0.002) | 0.54 (0.42,0.68) | <0.001 (<0.001) | 0.86 (0.57,1.3) | 0.466 (7.919) | 0.93 (0.72,1.21) | 0.582 (9.891) | ** | ** | ** | ** |
| GLP-1 inhibitors | 0.69 (0.36,1.34) | 0.276 (4.691) | 0.67 (0.36,1.22) | 0.191 (3.251) | 1.36 (0.53,3.5) | 0.518 (8.814) | 1.01 (0.53,1.94) | 0.967 (16.439) | ** | ** | ** | ** |
| Laxatives | 1.25 (1.19,1.31) | <0.001 (<0.001) | 1.85 (1.76,1.95) | <0.001 (<0.001) | 3.9 (3.57,4.26) | <0.001 (<0.001) | 1.46 (1.39,1.53) | <0.001 (<0.001) | 1.31 (0.85, 2.03) | 0.228 | 1.08 (0.75, 1.55) | 0.688 |
| Serotonin inhibitors | 1.84 (1.76,1.93) | <0.001 (<0.001) | 1.83 (1.74,1.92) | <0.001 (<0.001) | 3.85 (3.52,4.2) | <0.001 (<0.001) | 0.95 (0.91,0.99) | 0.029 (0.493) | 0.81 (0.63, 1.04) | 0.094 | 0.88 (0.71, 1.09) | 0.245 |
| Dopamin antagonists | 2.42 (1.95,3.0) | <0.001 (<0.001) | 2.18 (1.73,2.74) | <0.001 (<0.001) | 4.25 (3.12,5.79) | <0.001 (<0.001) | 0.79 (0.66,0.94) | 0.006 (0.11) | ** | ** | ** | ** |
| Benzodiazepines | 1.3 (1.22,1.39) | <0.001 (<0.001) | 1.61 (1.5,1.72) | <0.001 (<0.001) | 2.7 (2.43,3.01) | <0.001 (<0.001) | 1.22 (1.15,1.29) | <0.001 (<0.001) | 1.08 (0.49, 2.35) | 0.855 | 0.95 (0.51, 1.77) | 0.881 |
| Selective beta-2-adrenoreceptor agonists | 0.74 (0.7,0.78) | <0.001 (<0.001) | 0.71 (0.67,0.75) | <0.001 (<0.001) | 0.83 (0.74,0.93) | 0.001 (0.022) | 0.95 (0.89,1.01) | 0.091 (1.54) | 0.83 (0.6,1.15) | 0.258 | 0.82 (0.62,1.09) | 0.172 |

**Figure 1:**
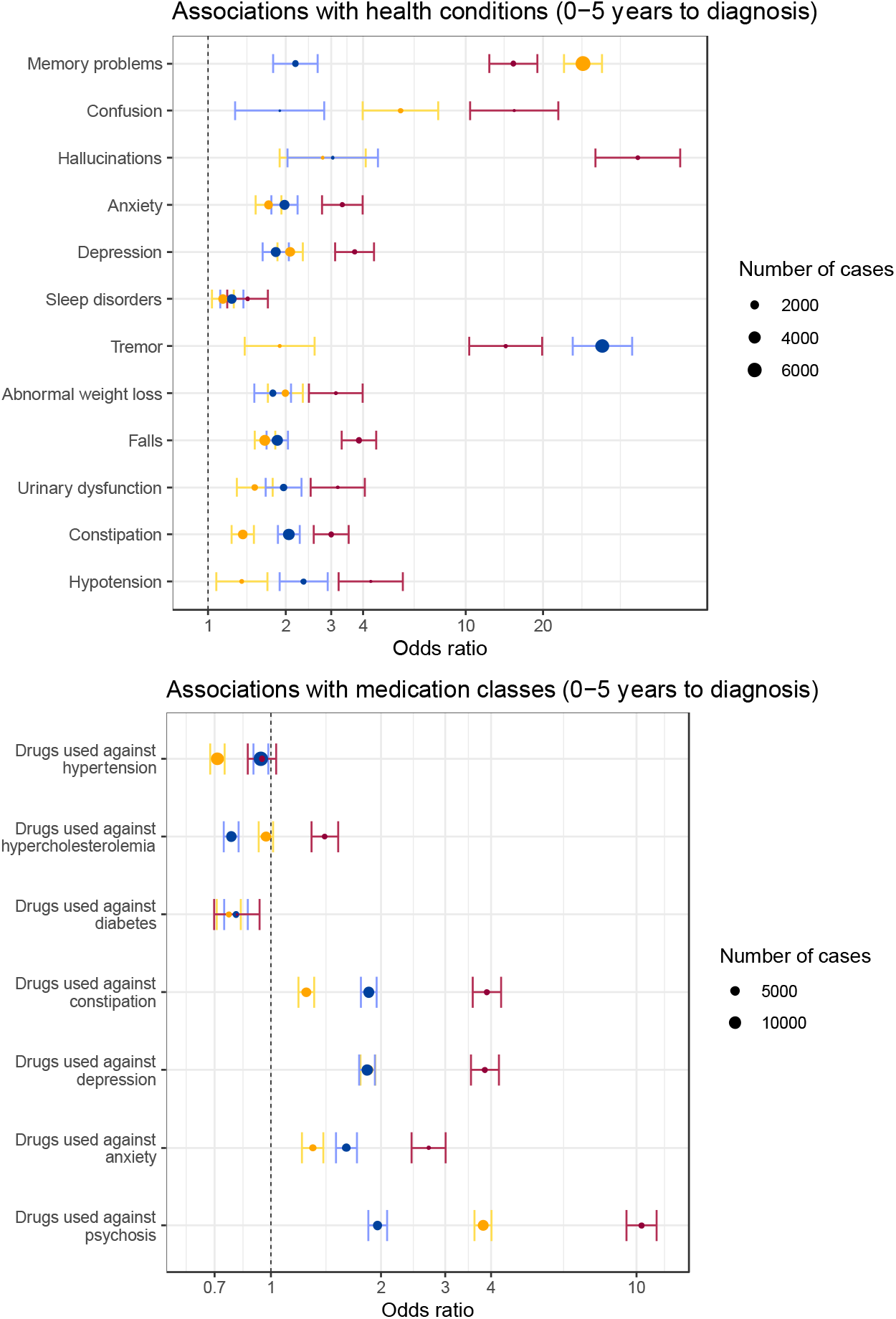
Disease specific odds ratio of multiple health conditions and classes of medications for Alzheimer’s disease (yellow), Parkinson’s disease (blue) and Lewy-bodies dementia (red) compared to controls. Odds ratios are corrected for sex and age at diagnosis. Odds ratios are computed in the time period between (0-5] years before a first diagnosis of the corresponding neurodegenerative disease. Confidence intervals are corrected for multiple comparisons.

**Figure 2:**
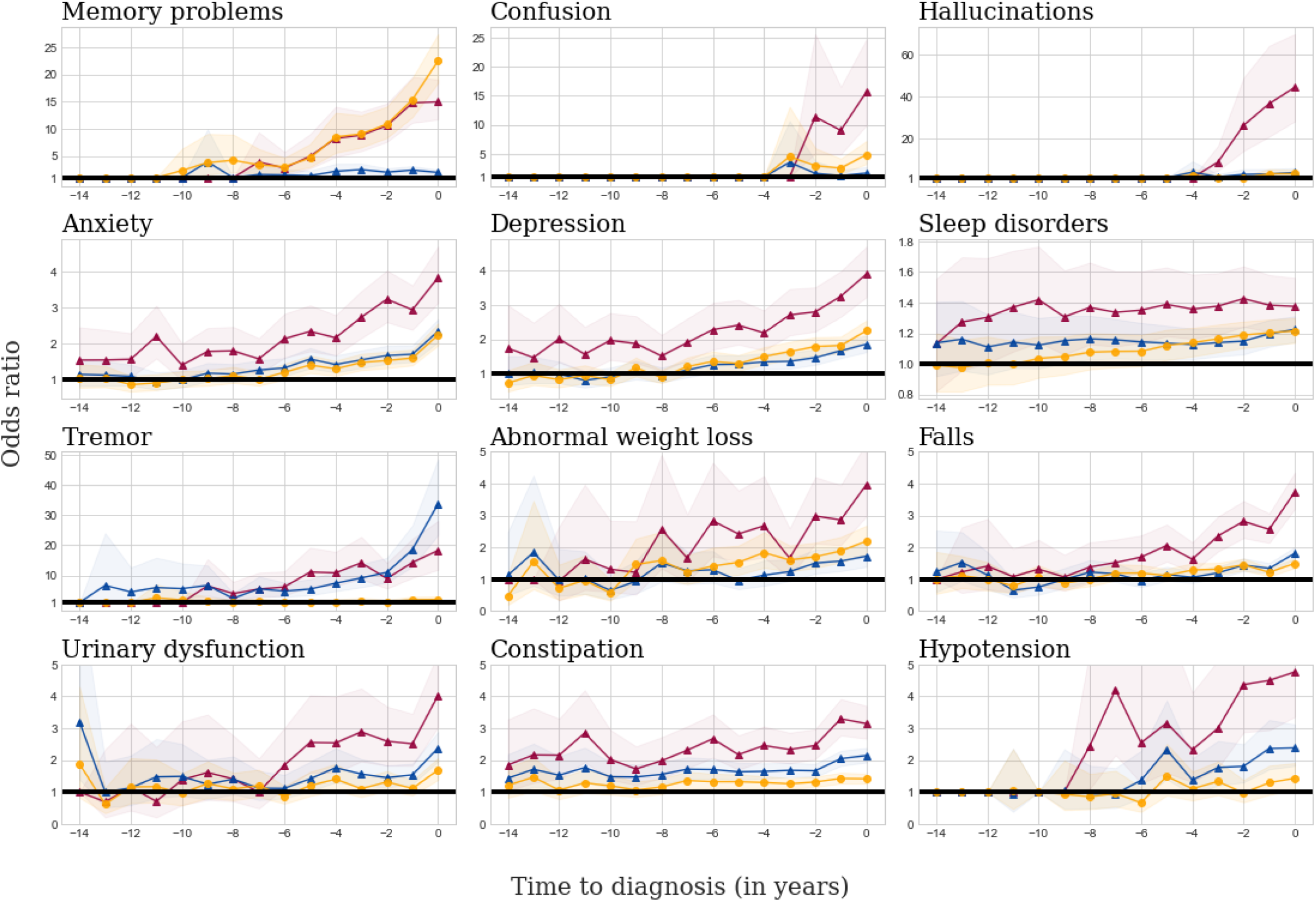
Disease specific odds ratio of mutlple health conditions for Alzheimer’s disease (yellow), Parkinson’s disease (blue) and Lewy-bodies dementia (red) compared to controls. Odds ratios are corrected for sex and age at diagnosis. The x-axis corresponds to the time (in years) before diagnosis. In the statististical analysis, we report odds ratios computed for following time period in years: [15,10), [10,5) and [5, diagnosis). Y-axis shows the odds ratio.

### Cross disorder analyses

The odds of the 12 health conditions before diagnosis was different when comparing between diseases (eTable 4). In the 5 years prior to diagnosis, memory problems were more frequent in AD compared to PD and DLB patients, while the constipation was more common before PD and DLB onset than before AD onset. When comparing PD and DLB, we found that tremor was more frequent in the years preceding PD diagnosis, whereas memory troubles, confusion, hallucinations, constipation, falls and depression were more frequent before the diagnosis of DLB. Laxatives were used more frequently prior to diagnosis of PD and DLB than of AD, in line with the higher prevalence of constipation in these conditions. (Table 4 and eTable 3). A higher beta-blocker agents prescription rate was observed in the years prior to PD and DLB diagnoses compared with AD patients. A higher prescription rate of statins was observed in the years prior to the DLB diagnosis while the opposite was observed prior to PD diagnosis, when compared with AD (Table 4). Similar results were obtained when restricting the PD population to patients with at least one prescription of antiparkinsonian drug (eFigure 4).

### Patients with Dementia with Lewy bodies

Patients with Lewy body dementia present stronger associations with most health conditions than AD and PD patients. DLB was strongly associated with depression, anxiety, autonomic dysfunction, constipation, urinary disorders and hypotension. The association with falls was also stronger in the DLB cohort than in the other neurodegenerative disorders (Figure 1). Neuroleptics were more frequently prescribed before DLB than AD and PD diagnoses, in accordance with their greater association with hallucination and confusion.

### Associations with the risk of developing dementia after a Parkinson’s disease diagnosis

To investigate whether the observed associations we describe above were specific to the disease or to the syndrome (dementia), we run a subsequent analysis in the subpopulation of PD patients, comparing PD patients who will develop a diagnosis of dementia and those who will remain free of dementia 5 years after PD diagnosis. The median age at diagnosis of PD was younger for PD patients who did not develop dementia in the five years after the first PD diagnosis (73 (65-79) years) than for PD patients who developed a dementia (79 (73-83) years) (eTable 7, p < 0.0001). There were more men than women in both groups (56% in the PD group without dementia and 59% in the PD group with dementia). When comparing the two groups, after correcting for sex and age at diagnosis, the prescriptions of drugs against hypercholesterolemia, constipation, depression and psychosis were individually associated with the development of a dementia in the five years after PD diagnosis (Figure 3). They all remained significantly associated with dementia in the multivariable model (Table 2). By contrast to what was found in the whole population, drugs against hypertension and anxiety were not individually associated with dementia after a Parkinson’s disease diagnosis.

**Figure 3:**
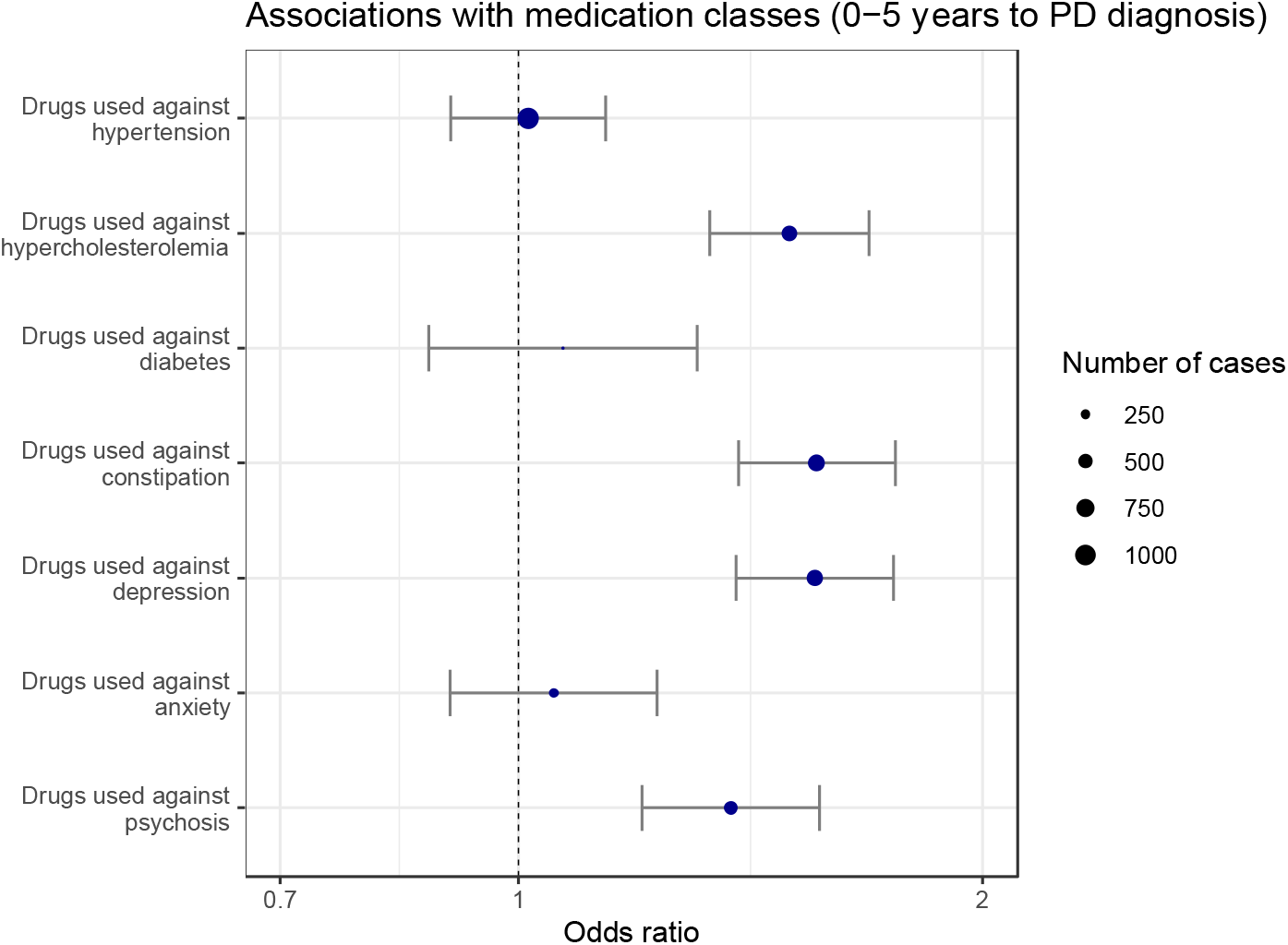
Disease specific odds ratio of multiple classes of medications for Parkinson’s patient with dementia in the five years following Parkinson’s disease diagnosis compared to Parkinson’s patients without dementia in the five years following Parkinson’s disease diagnosis. Odds ratios are corrected for sex and age at diagnosis. Odds ratios are computed in the time period between (0-5] years before a first Parkinson’s disease diagnosis. Confidence intervals are corrected for multiple comparisons.

### UKB replications

We used the UKB database to replicate the comparison in terms of medication usage between AD and PD. A lower prescription of statins was also observed in the years prior to diagnosis of PD as compared to AD (OR = 0.77, p= 0.01). As in the THIN database, there was no significant difference between AD and PD patients regarding agents acting on the renin– angiotensin–aldosterone system. When PD patients were compared with dementia cases (including vascular dementia and AD), prescriptions of all families of anti-hypertensives except diuretics (calcium channel blockers, beta blockers and agents acting on renin– angiotensin–aldosterone system) and statins (Table 4) were higher in individuals with subsequent dementia. We found that a higher prescription of statins was significantly associated with increased BMI, lower age at diagnosis and lower age at first visit in both the AD and PD cohorts but not significantly associated with any APOE genotypes (eTable13). The levels of LDL and HDL did not differ between PD and AD patients (eTable14), but we do not know if patients were cholesterol-lowering medications and the number of patients who had a blood sample before their diagnosis was very small (88 PD, 91 AD).

## Discussion

In this large cross-disorder analysis, we compared the prodromal symptoms and drug prescription before the diagnosis of three of the main neurodegenerative diseases (AD, PD and DLB). Our results complement previous approaches which tested seperately a range of prodromal features on PD^9^ or AD^8^, observable at the GP level. By constrasting AD with PD and DLB, we found cardinal motor or non-motor symptoms of each diseases to be more frequent than in controls at least 5 years before diagnosis: memory problems for AD, tremor for PD and hallucination, confusion, movement disorders and memory problems for DLB. We also observed that GP reported more falls in future DLB patients than in AD and PD patients in adequation with the McKeith 2017 criteria for DLB.^18^ We also found a robust association between all three neurological disorders and constipation in accordance with previous studies,^19^ even though the association was stronger for PD and DLB than for AD. More generally, we showed that all neurological disorders present more autonomic dysfunction in primary care compared to controls, including constipation, urinary disorders and hypotension several years before diagnosis, with a stronger association for PD and DLB than for AD. These autonomic dysfunctions might be used by GP in the future to better anticipate a neurodegenerative disease. This result may support the hypothesis that neurodegenerative diseases pathological mechanisms affect the periphery before spreading into the brain.^20^ These autonomic dysfunctions might alert the GP to a potential evolution towards a neurodegenerative disease.

We also investigated drug use and found a lower prescription of all the main antihypertensors families, including agents acting on the renin–angiotensin–aldosterone system (for all three diseases), calcium channel blockers (for AD and PD) and diuretics (for AD and DLB), in accordance with previous epidemiological studies showing an inverse association between antihypertensive drugs and AD, PD or DLB.^21^ These results may support the hypothesis that vascular diseases are a risk factor of neurodegenerative diseases^21^, a frequent comorbid condition of these diseases and that decreasing cardiovascular risks by prescribing antihypertensive drugs would help to decrease the risk of developing all neurodegenerative diseases. Alternatively, some of these drugs may have unspecific neuroprotective effects,^22^ as antidiabetics drugs that were less prescribed in the years preceding all three diseases.^23^ The design of this study did not allow us to distinguish between these two possibilities. The difference observed on beta-blocker agents which were more prescribed in individuals who later developed PD and DLB compared to AD or controls, confirms previous similar results and may be explained by their use to relieve tremor.^24^ The lower risk of developing PD with the use of Beta-2-adrenoreceptors and the higher risk with using beta-blocker agents may also be consistent with the current assumption that β2-adrenoreceptor is a regulator of the α-synuclein gene (*SNCA*).^25^ Benzodiazepines usage have been repeatedly linked to a higher risk of dementia^26^, and benzodiazepines were indeed more prescribed in the years preceding with all three diseases. Of note, all three diseases were also associated with increased rates of anxiety and sleep disorders. As above, the design of the study did not allow us to distinguish between a direct effect of benzodiazepines on risk of dementia or the consequence of the need to treat prodromal anxiety. Our observed association of benzodiazepines with PD and DLB suggests that this association is not specific to AD.

More surprisingly, we found a lower prescription of statins in years before the diagnosis of PD compared to dementia-related disorders (AD, DLB). These differences remained significant when adjusting for potential counfounding variable such as APOE status, BMI, education level and townsend deprivation status, and in the replication (UKB) dataset. This observation could have several explanations. First, since statins are prescribed in secondary prevention of cardiovascular diseases, it may result of higher vascular comorbidities in the AD and DLB groups.^27^ However, lipid lowering therapies intake was similar between AD and controls. Moreover we observed that anti-hypertensors were similarly prescribed in the three diseases, which suggests that the three populations may have similar vascular risk factors. Secondly, AD participants were screened out for vascular dementia in the UKB. Another explanation would be that subjects treated with lipid lowering therapies are at lower risk to develop PD. Indeed, higher cholesterol levels have been associated with a lower risk of PD,^28,29^ including a recent mendelian randomization analysis suggesting that this association could be causal.^30^ Finally, lipid lowering agents may have a protective effect on PD as supported by a study showing that the Mediterranean diet adherence is associated with a reduced risk of prodromal PD,^31^ even though similar studies also exist in AD.^32^ A recent randomized controlled trial of simvastatin for delay of disease progression in PD was negative,^33^ indicating that the association might not be causal. Another assumption would be that PD induces a global hypermetabolism and thus fat burning, which could indirectly lower cholesterol levels and thus a lower use of statins. The mechanisms by which lipid lowering therapies may be protective against PD remains to be understood.

Interestingly, the increased usage of statins prior the diagnosis of dementia in the whole population (i.e. AD or DLB) was also found when restricting the analysis in the PD population. Indeed, we found that PD patients who developed dementia within 5 years used more anti-cholesterol drugs at the pre-PD stage than patients not developing dementia. It may argue towards a specific role of statins in the development of dementia. Finally, we also identify other prescriptions associated with future development of dementia in the PD population (drugs against constipation and depression). Using past prescriptions of the general practitioner may be useful in the future to design approaches to distinguish PD patients who will develop dementia shortly after their first initial PD diagnosis.

This study is to our knowedge the largest addressing on the same database the prodromal phase of the three neurodegenerative diseases allowing to compare their clinical profile and treatment prescriptions as observed by general practitioners. Another notable strength of this study is the replication on the UK Biobank, with consistent results obtained for the two analysis. Our work has also several limitations. First, in the THIN database, the diagnosis of neurodegenerative diseases was made by general practitioners, which may result in a delayed diagnosis, explaining why we can observe diseases’ specific symptoms before their diagnosis. In the UKB database, diseases have to be detected during an hospitalisation or mentioned in death registers. Second, the frontiers of each complex neurodegenerative disease may not be defined perfectly, the zero one distinction being a simplification with respect to disorders heterogeneity^34^ and distinction between developing a dementia with Lewy Bodies or having at the same time PD and AD might be difficult to do in practice. However, the differences observed in the prodromal stage of DLB patients, show that DLB is not simply the addition of the symptoms of AD and PD at the GP level. Finally, in the THIN analysis, controls were matched with the AD population, which was older and had a higher proportion of female patients than the PD and DLB cohorts, and we may therefore have underestimated associations particularly in the younger PD population. AD patients in the UKB might also not be representative of the general patient population as they are still relatively young.

## Conclusion

Our results highlight the long-term clinical prodromal phase of neurodegenerative diseases, and therefore could enable more efficient primary and secondary prevention measures. To the best of our knowledge, this is the first study showing that there may be a differential role of lipid lowering therapies in the prodromal phase of neurodegenerative diseases. Further studies are needed to better understand the relationship between cholesterol levels, the risk of neurodegenerative diseases and the risk of developing dementia after a Parkinson’s disease diagnosis.

## Data Availability

The data used in the preparation of the article are available from the Cegedim company upon reasonable request.

## Contributors

TN, BCD, AS, JCC and SD conceived and designed the study. FM, LG, BCD, and ADB extracted the data. FM, LG and TN ensured data quality and TN, BCD, ADB, AS, JCC and SD analysed the data. TN and BCD generated the figures. TN, BCD, ADB, FM, LG, BL, NV, AS, SD and JCC interpreted the results and contributed to the writing of the final version of the manuscript. All authors agreed with the results and conclusions and approved the final draft.

## Acknowledgement

The research leading to these results has received funding from the joint program in neurodegenerative diseases (JPND) ANR-21-JPW2-0002-01 (LeMeReND) and the program “Investissements d’avenir” ANR-10-IAIHU-06. This work was funded in part by the French government under management of Agence Nationale de la Recherche as part of the “Investissements d’avenir” program, reference ANR-19-P3IA-0001 (PRAIRIE 3IA Institute). BCD is supported by a CJ Martin Fellowship (NHMRC, APP1161356) and the CNRS.

## Ethics committee approval

Approval was sorted before carrying out this study (and for the use of datasets). The study was a retrospective analysis of secondary anonymised patient data only. For the UK THIN database, data is only available to researchers carrying out approved medical research. Ethical approval was granted by the NHS South-East Multicentre Research Ethics Committee in 2003 (ref: 03/01/073) for establishment of the THIN database, and it was updated in 2011. A further update and approval was granted in 2020 by the NHS South Central, Oxford C Research Ethics Committee (Ref: 20/SC/0011). The study was approved by the THIN Scientific Research Committee (SRC) (SRC reference 20-002). Researchers willing to replicate the results should address a request to the Cegedim company. Informed consent was obtained from all UKB participants. Procedures are controlled by a dedicated Ethics and Guidance Council (http://www.UKBiobank.ac.uk/ethics), with the Ethics and Governance Framework available at http://www.UKBiobank.ac.uk/wp-content/uploads/2011/05/EGF20082.pdf. IRB approval was also obtained from the North West Multi-centre Research Ethics Committee. This research has been conducted using the UK Biobank Resource under Application Number 53185.

## Patient and Public involvement

We plan to disseminate the results of our research to the public and relevant patient community through the current publication and its related press release as well as presentations at scientific conferences.

The lead authors (AS, SD, JCC) affirm that this manuscript is an honest, accurate, and transparent account of the study being reported; that no important aspects of the study have been omitted; and that any discrepancies from the study as planned (and, if relevant, registered) have been explained.

